# Deep Learning-Based Single-View Echocardiographic Analysis for Prediction of Left Ventricular Outflow Tract Gradient Elevation After Transcatheter Aortic Valve Replacement

**DOI:** 10.64898/2026.03.27.26349567

**Authors:** Jee-Woong Choi, Jiesuck Park, Yeonyee E. Yoon, Jiyeon Kim, Jaeik Jeon, Yeonggul Jang, Seung-Ah Lee, Minjung Bak, Hong-Mi Choi, In-Chang Hwang, Goo-Yeong Cho

## Abstract

**Background and Objectives:** Dynamic left ventricular outflow tract (LVOT) gradient elevation can occur following transcatheter aortic valve replacement (TAVR) and remains difficult to predict with conventional transthoracic echocardiography. We aimed to determine whether a deep learning (DL) model, originally developed in hypertrophic cardiomyopathy (HCM) to predict severe LVOT obstruction (LVOTO) from resting 2D parasternal long-axis videos without Doppler inputs, could identify susceptibility to post-TAVR LVOT gradient elevation from pre-TAVR echocardiograms.

**Methods:** This retrospective study included 302 consecutive patients undergoing TAVR. The model was applied to pre-TAVR resting parasternal long-axis transthoracic echocardiography (TTE) videos to generate a patient-level DL index of LVOTO (DLi-LVOTO; range 0–100). Post-TAVR Doppler-defined LVOTO was defined as a peak pressure gradient ≥30 mmHg on follow-up TTE.

**Results:** Pre-TAVR LVOTO was present in 32 patients (10.6%) and post-TAVR LVOTO in 35 (11.6%). DLi-LVOTO was significantly elevated in patients who developed post-TAVR LVOTO (P<0.001). In multivariable analysis adjusting for conventional TTE parameters and pre-TAVR LVOTO status, DLi-LVOTO remained independently associated with post-TAVR LVOTO (adjusted OR 1.24, 95% CI 1.02–1.51 per 10-score increase, P=0.026), with an area under the receiver operating characteristic curve (AUROC) of 0.78 (95% CI 0.72–0.85). Among patients without pre-TAVR LVOTO, DLi-LVOTO retained independent predictive value (adjusted OR 1.61, 95% CI 1.27–2.14, P<0.001; AUROC 0.84, 95% CI 0.77–0.91).

**Conclusions:** A HCM-trained DL model predicts Doppler-defined post-TAVR LVOTO from pre-TAVR TTE, suggesting that DLi-LVOTO may reflect a shared obstructive phenotype beyond conventional structural assessments.

## INTRODUCTION

Severe aortic stenosis (AS) is increasingly treated with transcatheter aortic valve replacement (TAVR), particularly in older patients.^1^ TAVR effectively relieves fixed valvular obstruction and reduces left ventricular (LV) afterload; however, abrupt afterload reduction may unmask dynamic LV outflow tract (LVOT) gradient elevation in susceptible patients. The clinical significance of post-TAVR LVOT gradient elevation is heterogeneous, but clinically relevant hemodynamic compromise can occur in selected patients.^2–4^ Previous studies have suggested that AS-related remodeling features, including increased septal thickness, a small LV cavity, and a narrow LVOT, may predispose patients to dynamic obstruction after TAVR.^2,5^ However, these conventional transthoracic echocardiographic (TTE) parameters primarily represent static structural measurements and may not fully capture the combination of anatomical and dynamic features associated with post-TAVR LVOT gradient development. Identifying pre-procedural features associated with susceptibility to dynamic LVOT gradient elevation is therefore of interest, particularly for recognizing patients who may warrant closer post-procedural hemodynamic and echocardiographic assessment.

Recent advances in artificial intelligence have enabled deep learning (DL)-based analysis of TTE videos, allowing the extraction of structural and temporal information that may not be readily captured by conventional measurements.^6–9^ We previously developed and validated a DL model to predict severe LVOT obstruction (LVOTO) in patients with hypertrophic cardiomyopathy (HCM) using resting parasternal long-axis (PLAX) TTE videos without Doppler input.^6^ Although the model was originally trained in an HCM population, it integrates structural and dynamic information from the PLAX view, including features related to LV cavity geometry, the basal septum, LVOT, and mitral valve motion. Because some of these imaging features may also predispose patients with severe AS to dynamic obstruction after abrupt afterload relief, we hypothesized that the HCM-trained model might identify a shared obstructive imaging phenotype relevant to post-TAVR LVOT gradient elevation. Therefore, the present study aimed to investigate whether this HCM-trained DL model, applied to pre-TAVR resting PLAX TTE, could identify susceptibility to Doppler-defined LVOTO after TAVR in patients with severe AS.

## METHODS

### Study Population and Data Sources

We retrospectively analyzed consecutive patients who underwent TAVR for severe AS via the transfemoral approach at Seoul National University Bundang Hospital between February 2017 and July 2025. The primary inclusion criterion was the availability of both pre-TAVR and post-TAVR TTEs, with the TAVR date used as the index date. Pre-TAVR TTE was defined as echocardiography performed within 30 days prior to TAVR to reflect the most recent LV hemodynamic status. Post-TAVR TTE was defined as TTE performed between 7 and 180 days after TAVR. Post-TAVR TTE examinations performed within 1 week after TAVR for focused assessment of procedural complications were excluded. Among 351 eligible patients, 49 were excluded for the following reasons: absence of pre-TAVR echocardiography within 30 days before TAVR (n=6), absence of post-TAVR echocardiography between 7 and 180 days after TAVR (n=37), or valve-in-valve procedures (n=6). After applying the inclusion and exclusion criteria, 302 patients were included in the final analysis (**Figure 1**).

**Figure 1.**
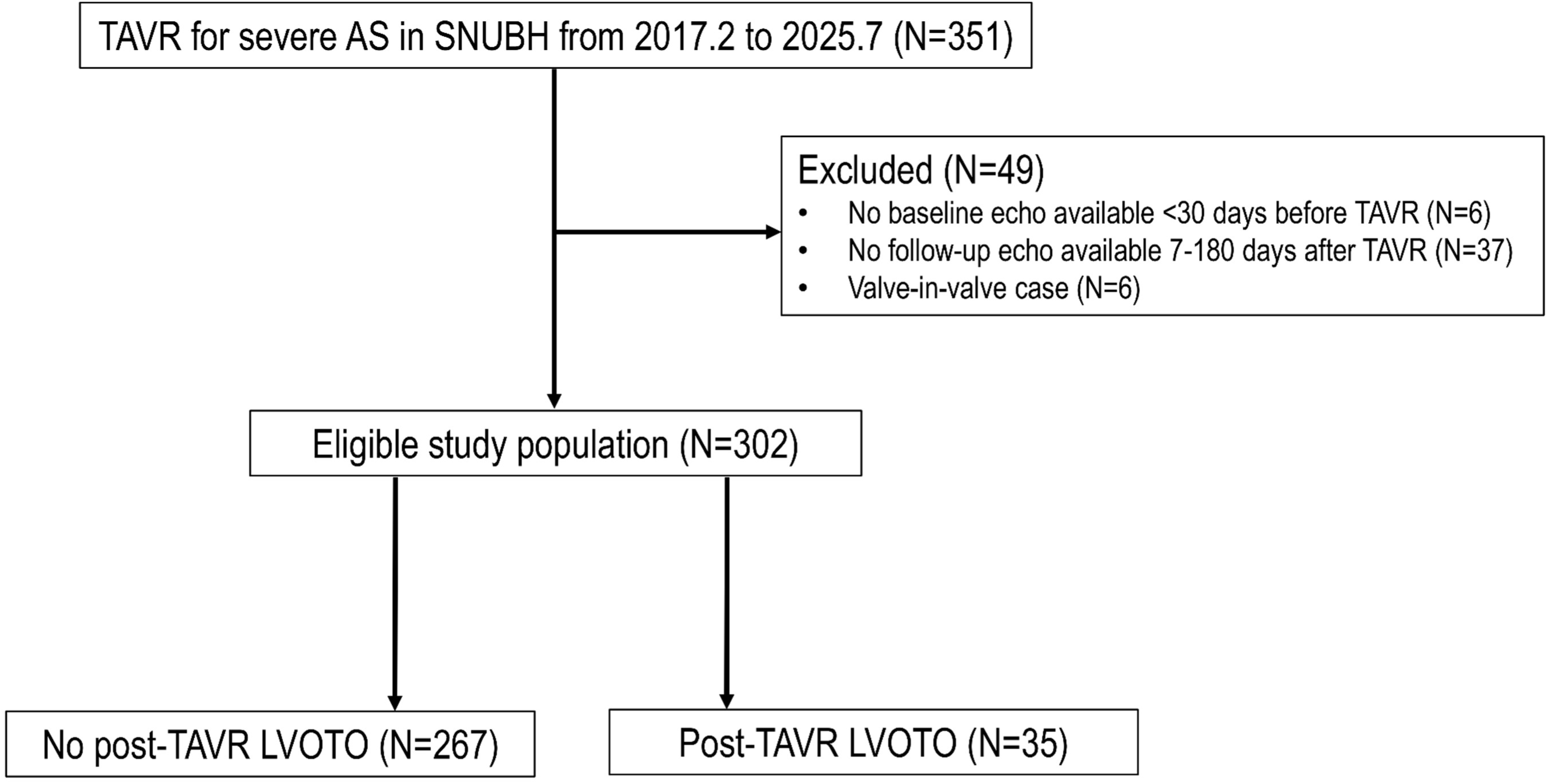
Study Flow Diagram. Of 351 patients who underwent TAVR for severe AS (February 2017–July 2025), 49 were excluded (absence of pre-TAVR TTE within 30 days [n=6], absence of post-TAVR TTE within 7–180 days [n=37], or valve-in-valve procedures [n=6]), yielding 302 patients for analysis. Post-TAVR LVOTO was identified in 35 patients (11.6%). Abbreviations: AS, aortic stenosis; LVOTO, left ventricular outflow tract obstruction; SNUBH, Seoul National University Bundang Hospital; TAVR, transcatheter aortic valve replacement; TTE, transthoracic echocardiography.

The study protocol complied with the Declaration of Helsinki and was approved by the Institutional Review Board of Seoul National University Bundang Hospital (approval number B-2602-1029-102). The requirement for informed consent was waived due to the retrospective design of the study.

### TTE Acquisition and Assessment of LVOTO

All TTE examinations were performed by trained echocardiographers or cardiologists and interpreted by board-certified cardiologists specialized in echocardiography, in accordance with contemporary guideline recommendations, as part of routine clinical care.^10^ Both pre- and post-TAVR examinations were standard resting TTE studies. Exercise or pharmacologic stress echocardiography was not included. However, provocation maneuvers routinely available during standard TTE, including the Valsalva maneuver, were systematically performed in patients with TTE features suggestive of LVOTO, such as hypercontractile LV, a sigmoid septum, or LVOTO on color Doppler. Baseline structural parameters were measured, including LV septal and posterior wall thickness, LV dimensions and volumes, and LVOT diameter.

For the assessment of LVOTO, continuous-wave Doppler interrogation was performed in the apical 3-chamber or 5-chamber view to measure LVOT peak velocity (V_max_, m/s). The peak PG across the LVOT was calculated using the simplified Bernoulli equation (PG = 4 × V_max_²). Provocation was not systematically performed in all patients or according to a uniform prespecified protocol. Rather, routine bedside provocation, primarily the Valsalva maneuver, was selectively performed when resting TTE demonstrated features suggestive of dynamic LVOTO, such as a hypercontractile LV, sigmoid septal morphology, or LVOT flow acceleration on color Doppler. The site of obstruction was confirmed at the LVOT level based on the location of flow acceleration and Doppler interrogation; midventricular or other intraventricular gradients were not classified as LVOTO. When provocation induced or increased the LVOT gradient, the highest peak PG obtained during the maneuver was used for classification; otherwise, the resting PG was used. For the present study, Doppler-defined LVOTO was operationally defined as an LVOT peak PG ≥ 30 mmHg using either the resting or provoked value, consistent with conventional HCM criteria.^11,12^ Resting LVOTO was defined as a peak LVOT PG ≥30 mmHg at rest, whereas provocation-induced LVOTO was defined as a peak LVOT PG ≥30 mmHg achieved only during provocation.

### DL Model for LVOTO prediction

The development and validation process of the DL model used in this study have been described previously.^6^ Briefly, the model was originally developed to predict severe LVOTO (peak PG ≥50 mmHg) in patients with HCM using single-view resting two-dimensional (2D) PLAX TTE videos without Doppler inputs. The backbone architecture is a modified R(2+1)D-18 network derived from ResNet-18, employing factorized three-dimensional convolutions (2D spatial + 1D temporal) to preserve full temporal resolution.^13,14^ To enhance motion sensitivity, the model incorporates automated M-mode generation via a spatial transformer network. The network learns an optimal M-mode line in a fully differentiable manner and extracts motion features along that trajectory. An auxiliary mean-squared-error loss constrains the learned trajectory to pass through the MV tip, thereby aligning the representation with clinically relevant anatomy. Multilevel M-mode features extracted at different network depths are fused to form comprehensive motion embeddings. Generated M-mode representations are processed with a pretrained EfficientNet-B3,^15,16^ and spatiotemporal (B-mode) features are then fused via concatenation. Thus, the model integrates both spatial/morphological and temporal/dynamic information from PLAX videos rather than relying on motion information alone. The model was trained using supervised learning with a primary binary classification objective for severe LVOTO (peak PG ≥50 mmHg), optimized by binary cross-entropy loss. Because LVOTO is defined by peak PG, often augmented by provocation, whereas input PLAX videos were obtained at rest, an auxiliary regression task was added to predict resting LVOT PG. This encouraged the network to extract subtle hemodynamic correlations from resting images. An early-exit strategy with intermediate auxiliary classifiers was implemented to improve gradient propagation and training stability. The total loss was the equally weighted sum of three components: anatomical constraint (mean squared error), LVOTO classification (binary cross-entropy), and PG regression (mean absolute error). All PLAX videos were normalized to 24 frames per second and resized to 224 × 224 pixels. Data augmentation techniques were applied to improve model robustness and included noise injection, sector masking, haze simulation, depth attenuation, dynamic gain variation, brightness and contrast adjustment, temporal perturbation, sharpening, and geometric transformations.

The model outputs a DL index of LVOTO (DLi-LVOTO; range: 0–100), a patient-level quantitative score that predicts the likelihood of LVOTO. In the present study, DLi-LVOTO was derived from pre-TAVR TTE data and used to predict post-TAVR LVOTO. Post-TAVR LVOTO assessment was based on clinical echocardiographic reports prior to and independent of DLi-LVOTO computation. When multiple PLAX clips were available for a patient, the final DLi-LVOTO value was calculated by averaging predictions across clips. This model is derived from the HCM analysis module (SoniX EchoPilot-CMP) of our AI-based TTE analysis solution (SONIX Health v 2.0; Ontact Health Co., Ltd.; Seoul, Republic of Korea), which integrates automated view classification and measurement functions.

### Statistical Methods

To evaluate the feasibility of applying DLi-LVOTO in patients with AS, DLi-LVOTO values were first compared by LVOTO status on pre-TAVR TTE. Distributions of DLi-LVOTO were visualized using box-and-whisker plots. The same analysis was then performed according to post-TAVR LVOTO status. To assess model interpretability, saliency maps were generated using Gradient-weighted Class Activation Mapping (Grad-CAM).^17^ Representative saliency maps were produced using pre-TAVR TTE images to highlight image regions that most strongly influenced LVOTO predictions.

Patients were categorized according to post-TAVR LVOTO status. Baseline clinical and TTE characteristics were compared between groups using the Kruskal-Wallis test for continuous variables and Fisher’s exact test or the chi-square test for categorical variables, as appropriate. To evaluate the association between DLi-LVOTO and post-TAVR LVOTO, univariate logistic regression analyses were performed. DLi-LVOTO was analyzed as a continuous variable. Additional clinical and TTE variables potentially associated with post-TAVR LVOTO were also evaluated, including age, sex, LV septal and posterior wall thickness, LV end-diastolic dimension and volume, LV ejection fraction, LVOT diameter, and the presence of sigmoid septum, mitral annular calcification, and pre-TAVR LVOTO. All candidate variables were entered into the initial multivariable model regardless of their univariate P-value. Given the relatively limited number of outcome events, multivariable analysis was performed using Firth’s penalized maximum likelihood logistic regression to reduce small-sample bias and potential separation. Backward elimination was then performed based on penalized likelihood ratio tests, with a retention criterion of P<0.20. To assess potential multicollinearity between DLi-LVOTO and covariates, the variance inflation factor (VIF) was calculated.

Receiver operating characteristic (ROC) curve analysis was performed to evaluate the discriminative performance of DLi-LVOTO for post-TAVR LVOTO by calculating the area under the ROC curve (AUROC), and the optimal cutoff was determined using Youden’s index. At the Youden-derived cutoff, sensitivity, specificity, positive predictive value (PPV), negative predictive value (NPV), positive and negative likelihood ratios (LR +, and LR -, respectively), and the proportion of patients classified above the cutoff were calculated. This internally derived cutoff was considered exploratory and requires external validation before clinical application. To assess model stability and account for the limited number of outcome events, the discriminative performance was internally validated using 1,000 bootstrap resamples, and a bootstrap-validated AUROC was additionally reported.

To evaluate the incremental predictive value of DLi-LVOTO beyond conventional echocardiographic parameters, three regression models were constructed and compared: (1) a conventional TTE model comprising significant univariable predictors (excluding DLi-LVOTO), (2) DLi-LVOTO alone, and (3) a combined model incorporating both. Differences in discrimination between the conventional and combined models were assessed using AUROC comparison, and the additional predictive value of DLi-LVOTO was further quantified using category-free net reclassification improvement (NRI).

As a sensitivity analysis, the independent and incremental predictive value of DLi-LVOTO was further evaluated in the subgroup of patients without pre-TAVR LVOTO to assess its ability to predict new-onset post-TAVR LVOTO.

A two-sided P value < 0.05 was considered statistically significant. All statistical analyses were performed using R software version 4.3.2 (R Foundation for Statistical Computing, Vienna, Austria).

## RESULTS

### Baseline Clinical and TTE Characteristics

Among the 302 patients included in the study population, the median age was 83 years (interquartile range [IQR] 80–86), and 135 (44.7%) were male. Pre-TAVR TTE was performed at a median of 10 days (IQR 1–24) before TAVR, and LVOTO was present in 32 patients (10.6%); of these, 5 (15.6%) had resting LVOTO and 27 (84.4%) had provocation-induced LVOTO. Post-TAVR TTE was performed at a median of 47 days (IQR 37–63) after TAVR, and LVOTO was identified in 35 patients (11.6%), of whom 16 (45.7%) demonstrated resting LVOTO and 19 (54.3%) had provocation-induced LVOTO. The majority of post-TAVR TTE studies were performed within 2 months after TAVR (216 of 302 patients, 71.5%). The prevalence of post-TAVR LVOTO was 10.2% at ≤2 months and 15.1% beyond 2 months, without a statistically significant difference according to follow-up timing.

When stratified by post-TAVR LVOTO status, baseline clinical characteristics were generally comparable between patients with and without post-TAVR LVOTO (**Table 1**). In contrast, several pre-TAVR TTE parameters differed between groups. Patients who developed post-TAVR LVOTO had smaller LV cavity size, reflected by lower LV end-diastolic dimension (LVEDD; 41 vs 45 mm, P=0.001) and LV end-diastolic volume (62 vs 79 mL, P=0.008). These patients also demonstrated higher LV ejection fraction (LVEF) (67% vs 62%, P=0.001). Left atrial (LA) size was also smaller in the LVOTO group, with lower LA volume index (46 vs. 54 mL/m², P=0.003). The distribution of the sigmoid septum and mitral annular calcification was similar between groups. However, patients in the post-TAVR LVOTO group had a smaller LVOT diameter (21 vs 22 mm, P=0.001) and received a smaller prosthesis (valve size: median 25 mm [IQR 23–26] vs. 26 mm [IQR 26–29], P<0.001). Additionally, pre-TAVR LVOTO was more prevalent among patients with post-TAVR LVOTO than among those without (57.1% vs 4.5%, P<0.001).

**Table 1.** Baseline characteristics.

| **Variable** | **Overall Population**<br>**(n=302)** | **No post-TAVR LVOTO**<br>**(N=267)** | **Post-TAVR LVOTO**<br>**(N=35)** | **p for diff** |
| --- | --- | --- | --- | --- |
| Age, years | 83 (80–86) | 83 (80–86) | 83 (79–86) | 0.362 |
| Male | 135 (44.7) | 122 (45.7) | 13 (37.1) | 0.451 |
| BMI, kg/m<sup>2</sup> | 24.3 (22.2–26.5) | 24.2 (22.2–26.6) | 24.5 (23.2–25.8) | 0.923 |
| SBP, mmHg | 130 (118–144) | 130 (117–143) | 132 (126–145) | 0.397 |
| DBP, mmHg | 66 (58–74) | 66 (58–73) | 67 (58–75) | 0.884 |
| **Procedural characteristics** |  |  |  |  |
| Valve type |  |  |  | 0.430 |
| Balloon-expandable | 107 (35.4) | 92 (34.5) | 15 (42.9) |  |
| Self-expanding | 195 (64.6) | 175 (65.5) | 20 (57.1) |  |
| Valve size, mm | 26 (25–29) | 26 (26–29) | 25 (23–26) | <0.001 |
| **Pre-TAVR TTE** |  |  |  |  |
| IVS, mm | 12 (11–14) | 12 (11–14) | 12 (11–14) | 0.852 |
| LVEDD, mm | 44 (41–48) | 45 (42–49) | 41 (38–45) | 0.001 |
| PW, mm | 11 (10–12) | 11 (10–12) | 12 (11–13) | 0.140 |
| LVEDV, ml | 77 (62–100) | 79 (64–102) | 62 (53–87) | 0.008 |
| LVEF, % | 63 (58–67) | 62 (57–66) | 67 (65–69) | 0.001 |
| LVMI, g/m2 | 119 (100–141) | 120 (101–141) | 109 (89–134) | 0.060 |
| LAVI, ml/m2 | 53 (41–67) | 54 (43–68) | 46 (38–57) | 0.003 |
| AV Vmax, m/s | 4.6 (4.3–5.1) | 4.6 (4.2–5.1) | 4.8 (4.5–5.3) | 0.065 |
| AV mPG, mmHg | 51 (42–63) | 50 (41–61) | 53 (47–67) | 0.099 |
| E/e' | 18 (14–23) | 18 (14–23) | 19 (14–25) | 0.639 |
| TR Vmax, m/s | 2.7 (2.5–3.0) | 2.7 (2.5–3.0) | 2.7 (2.4–3.0) | 0.649 |
| Sigmoid septum | 31 (10.3) | 24 (9.0) | 7 (20.0) | 0.558 |
| Mitral annular calcification | 26 (8.6) | 21 (7.9) | 5 (14.3) | 0.345 |
| LVOT diameter, mm | 22 (21–22) | 22 (21–23) | 21 (20–22) | 0.001 |
| DLi-LVOTO | 26 (9–54) | 18 (8–49) | 54 (40–74) | 0.001 |
| Pre-TAVR LVOTO | 32 (10.6) | 12 (4.5) | 20 (57.1) | <0.001 |

Data are presented as the median (interquartile range) for continuous variables and number (percentage) for categorical variables.
Abbreviations: AV, aortic valve; BMI, body mass index; DBP, diastolic blood pressure; DLi-LVOTO, deep learning index for left ventricular outflow tract obstruction; IVS, interventricular septum thickness; LAVI, left atrial volume index; LVEDD, left ventricular end-diastolic diameter; LVEDV, left ventricular end-diastolic volume; LVEF, left ventricular ejection fraction; LVMI, left ventricular mass index; LVOT, left ventricular outflow tract; LVOTO, left ventricular outflow tract obstruction; mPG, mean pressure gradient; PW, posterior wall thickness; SBP, systolic blood pressure; TAVR, transcatheter aortic valve replacement; TR, tricuspid regurgitation.; TTE, transthoracic echocardiography; Vmax, maximum velocity.

### Distribution of DLi-LVOTO According to Pre- and Post-TAVR LVOTO

DLi-LVOTO was computed from pre-TAVR PLAX TTE videos for all 302 patients. In the overall study population, the median DLi-LVOTO was 26 (IQR 9–54). DLi-LVOTO values were significantly higher in patients with pre-TAVR LVOTO compared with those without LVOTO (**Figure 2A**). This finding suggests that the DLi-LVOTO, originally developed in patients with HCM, captures structural and motion features associated with dynamic LVOTO even in patients with AS. When pre-TAVR DLi-LVOTO was examined according to the occurrence of post-TAVR LVOTO, patients who subsequently developed LVOTO showed markedly higher DLi-LVOTO values at the pre-TAVR stage than those who did not (**Figure 2B**). Overall, DLi-LVOTO demonstrated a clear rightward shift in patients with LVOTO at both pre- and post-TAVR stages, indicating that the DL-based index reflects the hemodynamic phenotype associated with dynamic LVOT obstruction.

**Figure 2.**
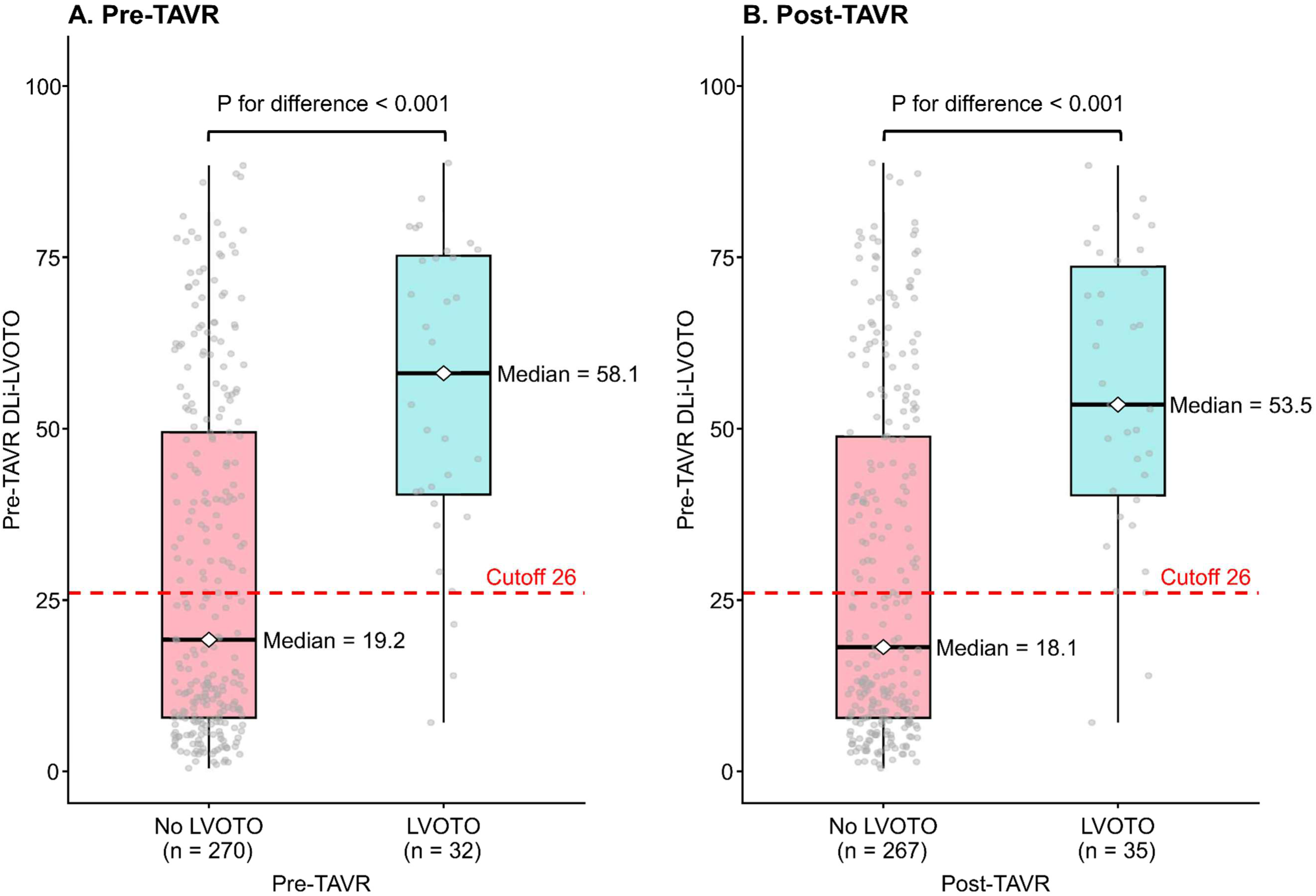
Distribution of Pre-TAVR DLi-LVOTO According to the presence of LVOTO at Pre- and Post-TAVR Stages. Box-and-whisker plots of pre-TAVR DLi-LVOTO, stratified by the presence of LVOTO on (A) pre-TAVR and (B) post-TAVR TTE. Pre-TAVR DLi-LVOTO was significantly higher in patients with LVOTO (resting or provocation-induced) at both timepoints (all P <0.001). Abbreviations: DLi-LVOTO, deep learning index for left ventricular outflow tract obstruction; LVOTO, left ventricular outflow tract obstruction; TAVR, transcatheter aortic valve replacement; TTE, transthoracic echocardiography.

### Prediction Performance of Pre-TAVR DLi-LVOTO for Post-TAVR LVOTO

In the overall study population, pre-TAVR TTE parameters were assessed for their association with post-TAVR LVOTO. In univariable analyses, several TTE variables were associated with post-TAVR LVOTO, including smaller LV cavity size and LV volume, higher LVEF, smaller LVOT diameter, and the presence of pre-TAVR LVOTO (**Table 2**). Higher pre-TAVR DLi-LVOTO values were also significantly associated with the occurrence of post-TAVR LVOTO (odds ratio [OR] 1.48, 95% CI 1.28–1.73 per 10-score increase, P<0.001). In the final multivariable model, pre-TAVR DLi-LVOTO remained independently associated with post-TAVR LVOTO (adjusted OR 1.24, 95% CI 1.02–1.51 per 10-score increase, P=0.026), along with pre-TAVR LVOTO (adjusted OR 11.00, 95% CI 4.16–30.88, P<0.001), posterior wall thickness (adjusted OR 1.38, 95% CI 1.01–1.92, P=0.040), and LVEF (adjusted OR 2.83, 95% CI 1.20–7.32 per 10% increase, P=0.016). The VIF for pre-TAVR DLi-LVOTO in the multivariable model was 1.48, indicating no significant multicollinearity with other covariates, including pre-TAVR LVOTO. To further examine the relationship between DLi-LVOTO and conventional echocardiographic features associated with dynamic obstruction, correlation analyses were performed. Correlation analysis demonstrated moderate associations of DLi-LVOTO with LV end-diastolic dimension (ρ = −0.50), LVEF (ρ = 0.39), LV end-diastolic volume (ρ = −0.38), and LVOT diameter (ρ = −0.32), whereas correlations with interventricular septal and posterior wall thickness were weak or negligible (**Supplementary Figure 1**). Pre-TAVR DLi-LVOTO demonstrated good discrimination performance for post-TAVR LVOTO (AUROC 0.78, 95% CI 0.72–0.85; bootstrap-validated AUROC 0.79, 95% CI 0.71–0.85) (**Figure 3A**). The optimal cutoff value determined by Youden’s index was 26, yielding a sensitivity of 94.3%, specificity of 56.9%, PPV of 22.3%, NPV of 98.7%, LR+ of 2.19, and LR− of 0.10. At this threshold, 49.0% of the overall population was classified as high risk. This internally derived cutoff should be considered exploratory and requires external validation before clinical application. To evaluate the incremental predictive value of DLi-LVOTO beyond conventional echocardiographic parameters, three models were compared in the overall population (**Table 3**). The conventional TTE model achieved an AUROC of 0.90 (95% CI 0.85–0.95). The addition of DLi-LVOTO demonstrated modest improvement in model discrimination (combined model AUROC 0.92, 95% CI 0.88–0.96; ΔAUROC +0.02, 95% CI 0.00–0.03, P=0.044; category-free NRI +0.67, P=0.014). Calibration of the combined model was further assessed using 1,000 bootstrap resamples. The optimism-corrected calibration slope was 0.84, the calibration intercept was approximately 0, the Brier score was 0.075, and the optimism-corrected C-statistic was 0.893 (**Supplementary Figure 2**).

**Table 2.**
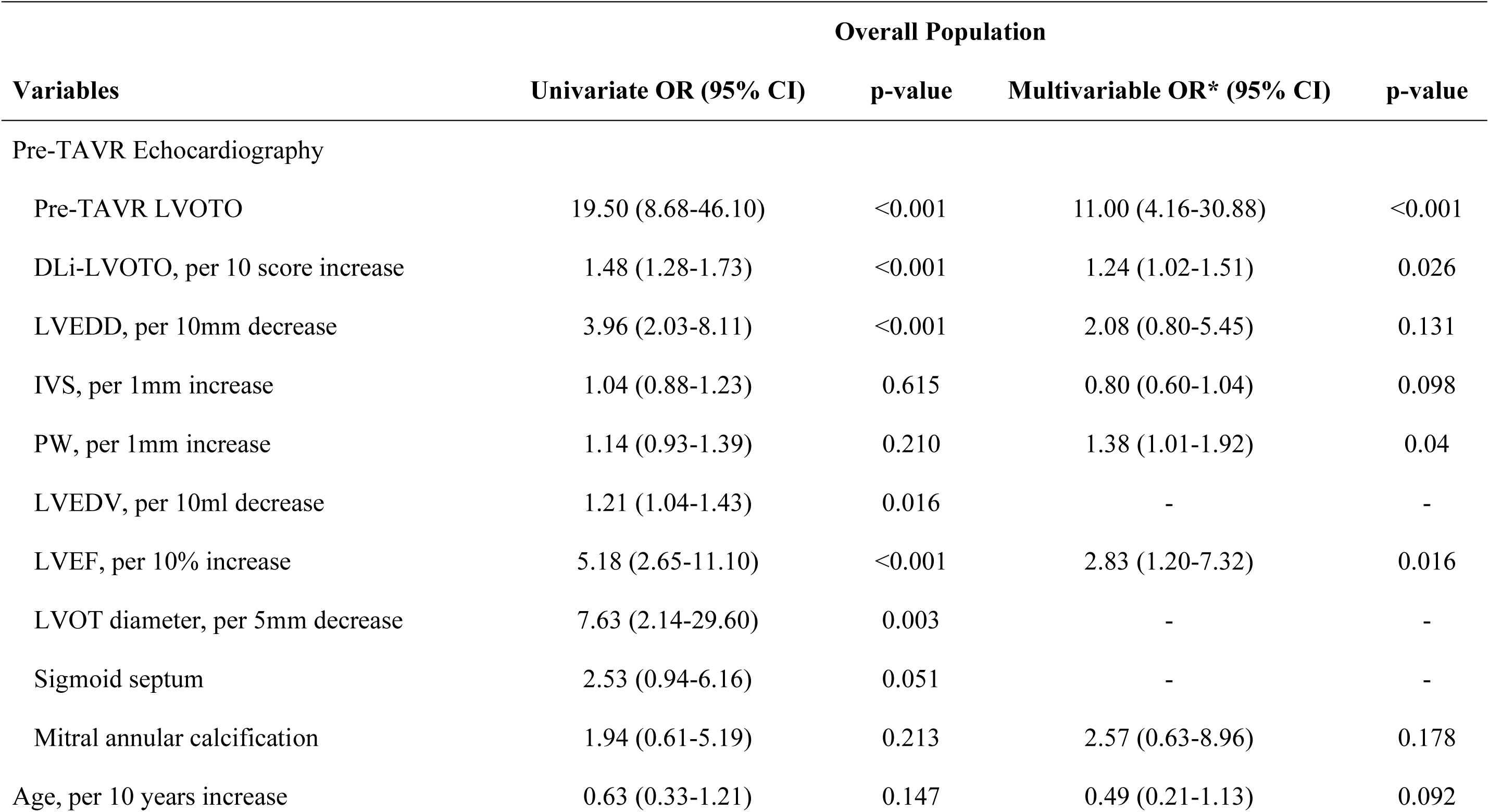
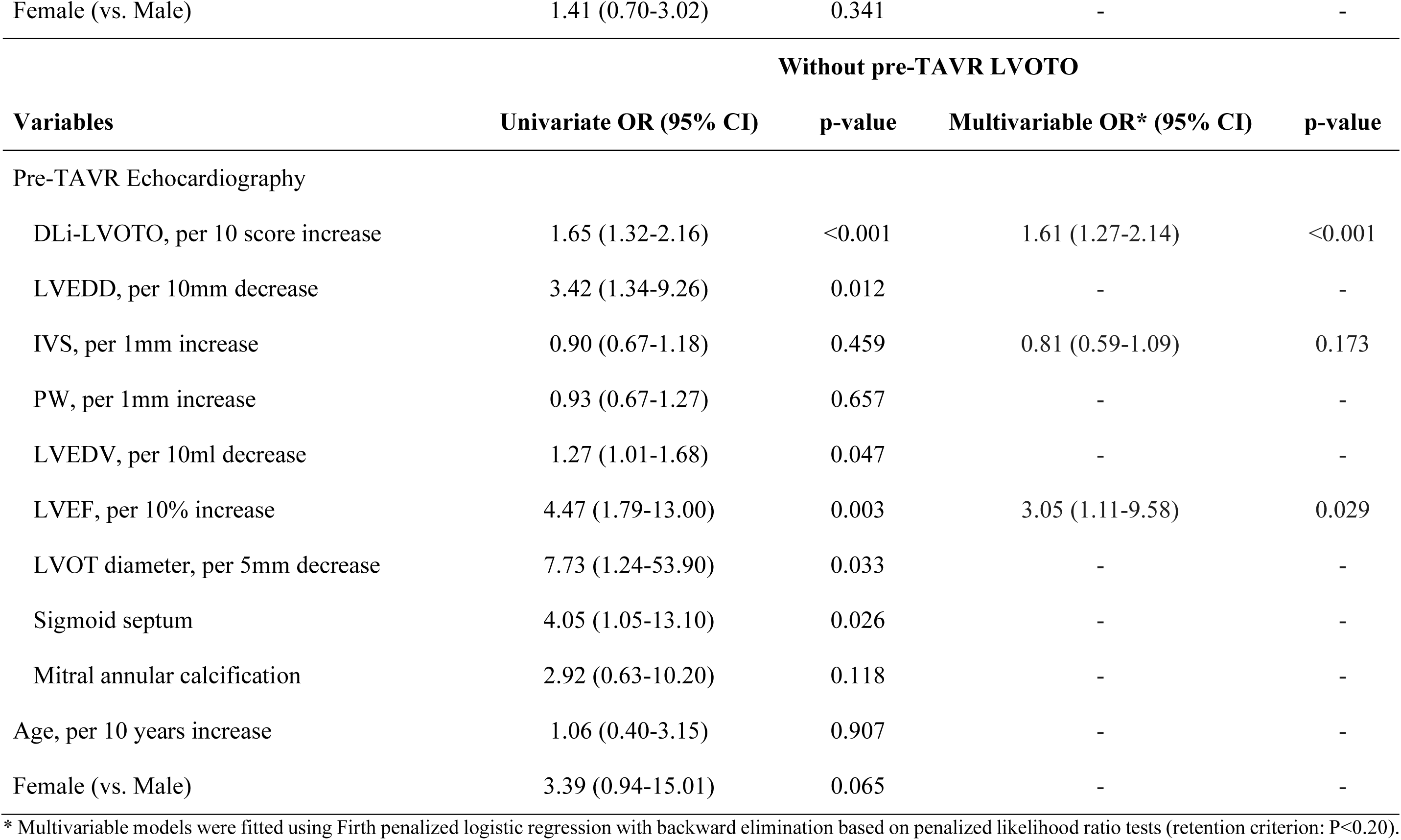
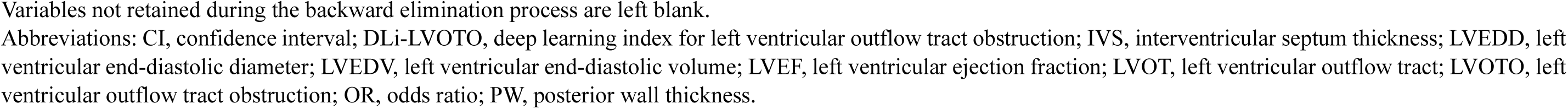
Association of pre-TAVR clinical and echocardiographic parameters with post-TAVR LVOTO.

**Figure 3.**
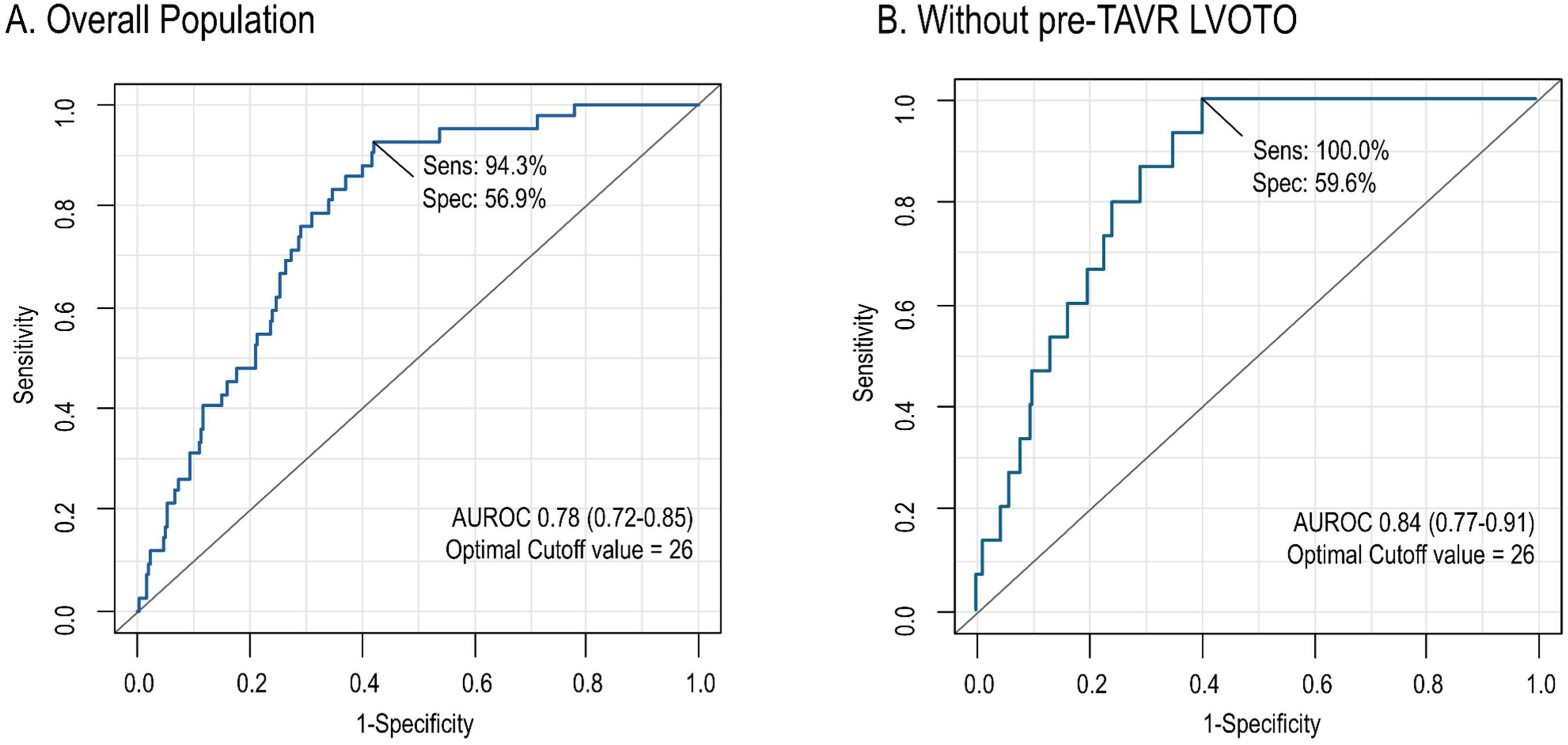
Discriminative Performance of Pre-TAVR DLi-LVOTO for Post-TAVR LVOTO. ROC curves for pre-TAVR DLi-LVOTO in (A) the overall population and (B) patients without pre-TAVR LVOTO. In the overall population, the AUROC was 0.78 (95% CI 0.72–0.85), and the optimal cutoff of 26 yielded a sensitivity of 94.3% and specificity of 56.9%. In patients without pre-TAVR LVOTO, the AUROC was 0.84 (95% CI 0.77–0.91), and the same optimal cutoff (26) yielded a sensitivity of 100.0% and specificity of 59.6%. Abbreviations: AUROC, area under the receiver operating characteristic curve; DLi-LVOTO, deep learning index for left ventricular outflow tract obstruction; LVOTO, left ventricular outflow tract obstruction; ROC, receiver operating characteristic; TAVR, transcatheter aortic valve replacement.

**Table 3.** Incremental Predictive Value of Pre-TAVR DLi-LVOTO Beyond Conventional Echocardiographic Parameters for Post-TAVR LVOTO.

|  | Conventional TTE model*<br>AUROC (95% CI) | Combined model†<br>AUROC (95% CI) | ΔAUROC | P value | NRI<br>(95% CI) | P<br>value |
| --- | --- | --- | --- | --- | --- | --- |
| Overall Population | 0.90 (0.85–0.95) | 0.92 (0.88–0.96) | +0.02<br>(0.00–0.03) | 0.044 | +0.67<br>(0.16–0.98) | 0.014 |
| Without pre-TAVR LVOTO | 0.83 (0.73–0.92) | 0.88 (0.80–0.96) | +0.05<br>(0.00–0.10) | 0.033 | +0.98<br>(0.53–1.54) | <0.001 |

\* The conventional TTE model included the variables retained in the final multivariable model, excluding DLi-LVOTO (pre-TAVR LVOTO status, left ventricular end-diastolic dimension, interventricular septum thickness, posterior wall thickness, left ventricular ejection fraction, mitral annular calcification, and age).
† Combined model incorporated all variables of the conventional TTE model plus DLi-LVOTO.
Abbreviations: AUROC, area under the receiver operating characteristic curve; DLi-LVOTO, deep learning index for left ventricular outflow tract obstruction; LVOTO, left ventricular outflow tract obstruction; NRI, net reclassification improvement; TAVR, transcatheter aortic valve replacement; TTE, transthoracic echocardiography.

Given the variability in post-TAVR TTE timing, a sensitivity analyses were performed according to follow-up interval. The distribution of post-TAVR TTE timing, LVOTO prevalence, and peak LVOT gradients across 7–30, 31–60, 61–90, and 91–180 days is shown in **Supplementary Table 1**. Post-TAVR LVOTO occurred in 3 of 15 patients (20.0%), 19 of 201 (9.5%), 8 of 66 (12.1%), and 5 of 20 (25.0%) across these respective intervals. Among patients with LVOTO, median peak LVOT PGs were 51.8, 31.8, 41.0, and 36.0 mmHg, respectively, with no significant difference across intervals (P=0.062). Seven patients had a peak LVOT PG ≥50 mmHg, with no significant trend across follow-up intervals (P=0.229). When the analysis was restricted to narrower follow-up windows, the association between pre-TAVR DLi-LVOTO and post-TAVR LVOTO remained similar to that observed in the full cohort (31–90 days: OR 1.47 per 10-score increase, AUROC 0.787; 31–60 days: OR 1.45, AUROC 0.783; full cohort: OR 1.48, AUROC 0.784; **Supplementary Table 2**).

Sensitivity analyses were also performed according to LVOTO subtype, provocation performance, and severity. When the endpoint was restricted to resting LVOTO, thereby excluding provocation-induced events, pre-TAVR DLi-LVOTO remained associated with post-TAVR LVOTO (OR 1.37 per 10-score increase, 95% CI 1.13–1.69, P=0.002; AUROC 0.752). Pre-TAVR DLi-LVOTO was also associated with the performance of provocation during post-TAVR TTE (OR 1.46 per 10-score increase, 95% CI 1.27–1.67, P<0.001; AUROC 0.778). Similarly, when severe LVOTO was defined using a peak LVOT PG ≥50 mmHg, the association remained directionally consistent despite the limited number of events (n=7; OR 1.33, 95% CI 1.01–1.74, P=0.047; AUROC 0.743) (**Supplementary Table 2**). The distribution of DLi-LVOTO according to no LVOTO, resting LVOTO, and provocation-induced LVOTO is shown in **Supplementary Figure 3**.

Because pre-TAVR LVOTO was a strong predictor of post-TAVR LVOTO, additional sensitivity analysis was performed in patients without pre-TAVR LVOTO. In this subgroup, post-TAVR LVOTO occurred in 15 patients (5.6%). Pre-TAVR DLi-LVOTO remained independently associated with post-TAVR LVOTO in the final multivariable model (adjusted OR 1.61, 95% CI 1.27–2.14, P<0.001). Pre-TAVR DLi-LVOTO consistently showed good discrimination for post-TAVR LVOTO in this subgroup (AUROC 0.84, 95% CI 0.77–0.91; bootstrap-validated AUROC 0.84, 95% CI 0.76–0.91) (**Figure 3B**). The optimal cutoff value for predicting post-TAVR LVOTO was identical to that of the overall population (26), yielding a sensitivity of 100.0%, specificity of 59.6%, PPV of 12.6%, NPV of 100%, LR + of 2.45, and LR – of 0.00: 44.1% of patients in this subgroup were classified as high risk at this threshold. The incremental value of DLi-LVOTO beyond conventional TTE parameters was more pronounced in this subgroup (**Table 3**): the conventional TTE model achieved an AUROC of 0.83 (95% CI 0.73–0.92), which improved significantly with the addition of DLi-LVOTO (AUROC 0.88, 95% CI 0.80–0.96; ΔAUROC +0.05, 95% CI 0.00–0.10, P=0.033; category-free NRI +0.98, P<0.001).

### Representative Cases and Model Interpretability

Representative cases illustrating model predictions and interpretability are shown in **Figure 4** and **Supplemental Videos 1** and **2**. Case A was a woman in her seventies with significant LVOTO on pre-TAVR TTE (peak PG 38 mmHg). Pre-TAVR TTE showed an LVEDD of 46 mm and LVEF of 68%, and pre-TAVR DLi-LVOTO was high (DLi-LVOTO = 70). Grad-CAM saliency maps highlighted activation around the LVOT and anterior mitral leaflet—key anatomical structures involved in dynamic LVOT obstruction. After TAVR, LVEDD decreased to 37 mm, with a pronounced worsening of LVOTO (peak PG 74 mmHg).

**Figure 4.**
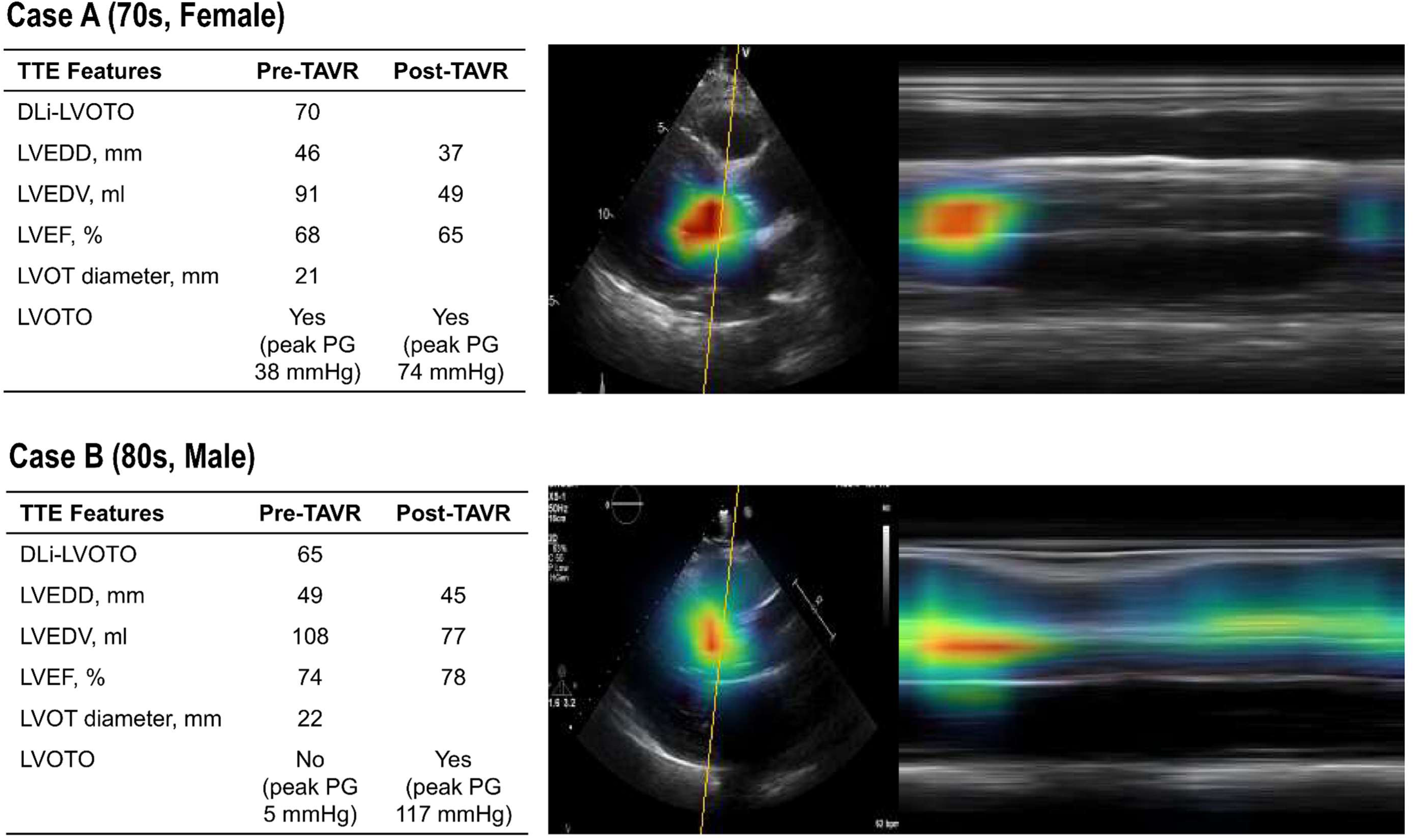
Representative Cases and Model Interpretability Analysis. Grad-CAM saliency maps and TTE features for two representative cases. Case A: a woman in her seventies with pre-TAVR LVOTO (peak PG 38 mmHg), LVEDD of 46 mm, LVEF of 68%, and pre-TAVR DLi-LVOTO of 70, who exhibited a pronounced worsening of LVOTO after TAVR (peak PG 74 mmHg). Case B: a male patient in his eighties without pre-TAVR LVOTO (peak PG 5 mmHg), with LVEDD of 49 mm and LVEF of 74%, but with an elevated pre-TAVR DLi-LVOTO of 65, who developed marked LVOTO after TAVR (peak PG 117 mmHg). In both cases, Grad-CAM saliency maps demonstrated activation in the LVOT and anterior mitral leaflet region, highlighting anatomically relevant features associated with post-TAVR LVOTO. Abbreviations: DLi-LVOTO, deep learning index for left ventricular outflow tract obstruction; Grad-CAM, Gradient-weighted Class Activation Mapping; LVEDD, left ventricular end-diastolic diameter; LVEDV, left ventricular end-diastolic volume; LVEF, left ventricular ejection fraction; LVOT, left ventricular outflow tract; LVOTO, left ventricular outflow tract obstruction; PG, pressure gradient; TAVR, transcatheter aortic valve replacement.

Case B was a male patient in his eighties. Pre-TAVR TTE showed an LVEDD of 49 mm and LVEF of 74%, without documented LVOTO (peak PG 5mmHg). Nevertheless, DLi-LVOTO derived from pre-TAVR TTE was high (DLi-LVOTO = 65). Grad-CAM saliency maps similarly demonstrated activation in the LVOT region. After TAVR, LVEDD remained at 45 mm; however, the patient developed a marked worsening of LVOTO, with a peak PG of 117 mmHg. Notably, Pre-TAVR DLi-LVOTO was elevated despite the absence of conventional markers of LVOTO risk, such as a small LV cavity or pre-TAVR LVOTO, suggesting that the DL model captures structural and dynamic features not readily identified by conventional echocardiographic assessment.

## DISCUSSION

This study demonstrated that an HCM-trained DL model applied to pre-TAVR PLAX TTE was independently associated with Doppler-defined LVOTO after TAVR in patients with severe AS. This association persisted after adjustment for conventional TTE parameters and pre-TAVR LVOTO status and was also observed in patients without pre-existing LVOTO. These findings support the feasibility of cross-domain application of a DLi-LVOTO and suggest that pre-TAVR PLAX imaging may contain features associated with susceptibility to dynamic LVOT gradient elevation after relief of valvular obstruction.

### Cross-domain Application of the DL Model for LVOTO Assessment

The rationale for applying an HCM-trained model to patients with severe AS is based not on an assumption that HCM-related LVOTO and post-TAVR LVOT gradient elevation represent the same physiological entity, but on the possibility that they share imaging features predisposing to dynamic obstruction. The PLAX view simultaneously depicts the LV cavity, basal septum, LVOT, mitral valve apparatus, and their systolic interactions, while also providing information on contractile behavior.^18^ The DL model integrates these structural and dynamic features from resting PLAX videos without Doppler input.^6^ In patients with severe AS, features such as LV hypertrophy, a relatively small LV cavity, narrow LVOT geometry, and hyperdynamic contraction may be present before TAVR and may predispose to gradient development after abrupt relief of valvular afterload. Accordingly, we interpret DLi-LVOTO in the present study as reflecting a PLAX-derived obstructive imaging phenotype rather than HCM-specific physiology or a direct measure of post-TAVR hemodynamics. The observation that DLi-LVOTO was higher in patients with pre-TAVR LVOTO and in those who subsequently developed post-TAVR LVOTO supports the presence of shared imaging features associated with dynamic obstruction across these disease settings. However, the actual development and magnitude of post-TAVR LVOT gradients are also influenced by loading conditions, contractility, heart rate, and pharmacologic factors that are not captured by the pre-TAVR PLAX video.

Previous studies further underscore the heterogeneous and dynamic nature of LV pressure gradients after TAVR. In a serial echocardiographic study by Tsuruta et al., post-TAVR LVOTO was most frequently observed at 3 months and decreased thereafter; notably, most cases represented midventricular obstruction rather than classic systolic anterior motion of MV (SAM)-related LVOTO, and the presence of obstruction was not associated with worse midterm clinical outcomes.^19^ More recent data similarly suggest that post-TAVR LVOTO is uncommon and has not been clearly associated with mortality or heart failure events.^20^ Nevertheless, isolated reports demonstrate that true SAM-related LVOTO can occasionally cause severe mitral regurgitation and acute hemodynamic compromise after TAVR.^21^

### Association of Pre-TAVR DLi-LVOTO with Post-TAVR LVOTO

Pre-TAVR LVOTO was present in 10.6% of patients and was the strongest independent predictor of post-TAVR LVOTO in multivariable analysis, consistent with prior reports identifying pre-existing dynamic obstruction as a major risk factor after afterload reduction.^2,5^ Importantly, pre-TAVR DLi-LVOTO remained independently associated with post-TAVR LVOTO even after adjustment for pre-TAVR LVOTO status, indicating that it captures predictive information beyond the binary presence or absence of pre-existing obstruction. No significant multicollinearity was observed, supporting the independent contribution of both variables to the model. The additive value of pre-TAVR DLi-LVOTO was particularly evident in patients without LVOTO at pre-TAVR stages, where the model maintained strong discriminative performance, suggesting that the index contains information associated with susceptibility to post-TAVR gradient development beyond the binary presence or absence of pre-existing LVOTO.

The association was also consistent across several sensitivity analyses addressing the heterogeneity of the post-TAVR endpoint. Restricting the analysis to narrower follow-up windows yielded effect estimates and discrimination similar to those of the full cohort. Importantly, the association persisted when the endpoint was restricted to resting LVOTO, thereby excluding provocation-induced events, and remained directionally consistent using the more stringent peak LVOT PG ≥50 mmHg threshold, although only seven patients met this criterion. These findings support the robustness of the observed association across alternative endpoint definitions, while the limited event counts, particularly for severe LVOTO, warrant cautious interpretation.

### Incremental Predictive Value of Pre-TAVR DLi-LVOTO Beyond Conventional TTE Parameters

Several pre-TAVR TTE parameters differed between patients with and without post-TAVR LVOTO, including smaller LV cavity size and volume, higher LVEF, smaller LVOT diameter, and smaller valve size.^5^ In contrast, the interventricular septal thickness did not differ significantly between groups. Although basal septal hypertrophy is classically cited as a risk factor for LVOTO,^5,18^ its discriminative power may be attenuated in this cohort, where virtually all patients already exhibited substantial septal thickening as a consequence of longstanding AS-related pressure overload. Similarly, LV mass index tended to be lower in the post-TAVR LVOTO group, which may appear counterintuitive. However, this likely reflects the denominator effect of indexing: patients in the post-TAVR LVOTO group had substantially smaller LV cavity dimensions, resulting in lower indexed mass despite comparable absolute wall thickness. These observations highlight the limitations of individual geometric parameters in characterizing the combination of structural and dynamic features associated with post-TAVR LVOT gradient development and support the potential complementary value of DLi-LVOTO.

DLi-LVOTO was associated with several conventional features linked to dynamic LVOT obstruction, including smaller LV cavity size, higher LVEF, and a smaller LVOT diameter. However, these correlations were only moderate, and no single conventional echocardiographic parameter appeared to account for the index. These findings suggest that DLi-LVOTO may reflect an integrated morphological and functional imaging phenotype rather than a distinct latent hemodynamic mechanism. The addition of DLi-LVOTO to the conventional TTE model resulted in a modest improvement in discrimination in the overall cohort, with a greater incremental improvement observed in patients without pre-TAVR LVOTO. Although bootstrap-based internal validation suggested limited optimism and generally adequate calibration of the combined model, these statistical improvements should not be interpreted as evidence of established clinical utility in the absence of external validation and clinically meaningful outcome assessment.

### Potential Clinical Implications and Future Directions

The present findings are primarily hypothesis-generating and should not yet be interpreted as supporting DLi-LVOTO-guided treatment or periprocedural management. Rather, they suggest that routinely acquired pre-TAVR PLAX videos may contain information associated with susceptibility to dynamic LVOT gradient elevation after TAVR. Because DLi-LVOTO requires only a routinely obtained PLAX video, this approach may provide a practical imaging strategy for further prospective evaluation. The internally derived cutoff of 26 showed high sensitivity and NPV but only modest specificity and low PPV, with nearly half of the cohort being classified as high risk. Accordingly, this cutoff should be regarded as exploratory rather than as a clinically validated action threshold and requires external validation before clinical application. Future studies should determine whether this imaging phenotype is reproducible under standardized post-TAVR conditions, associated with persistent or clinically consequential obstruction, and capable of providing incremental value for predicting clinically relevant outcomes.

### Study limitations

Several limitations of this study should be acknowledged. First, the present study is primarily hypothesis-generating, and the predictive value of DLi-LVOTO for post-TAVR LVOTO requires confirmation in adequately powered prospective studies before it can be considered for clinical implementation. Second, the present study evaluated the cross-domain application of a pre-existing HCM-trained model rather than developing or fine-tuning a TAVR-specific model. Although shared structural and dynamic imaging features associated with obstruction provided a rationale for this approach, a model specifically trained in patients undergoing TAVR may further improve predictive performance. The current cohort, with a limited number of post-TAVR LVOTO events, was not designed for de novo DL model development. Future studies using larger multicenter TAVR cohorts should determine whether TAVR-specific training or fine-tuning provides incremental value over the current cross-domain approach. Third, this was a retrospective study and the study population consisted exclusively of Korean patients from a single institution, which may limit generalizability to other ethnic groups and healthcare systems; application and validation in multi-institutional TAVR registries would further strengthen the external validity of DLi-LVOTO. Fourth, post-TAVR TTE was performed at varying timepoints within 180 days, and post-TAVR LVOT gradients may vary over time. Although additional analyses were performed according to follow-up timing, the present study cannot establish the persistence or temporal stability of Doppler-defined LVOTO. Fifth, provocation was performed selectively rather than systematically in all patients and was not conducted according to a uniform prespecified protocol. Higher pre-TAVR DLi-LVOTO was associated with a greater likelihood of undergoing provocation during post-TAVR TTE, supporting the possibility of differential ascertainment of provocation-induced LVOTO. However, the association between DLi-LVOTO and post-TAVR LVOTO persisted in the sensitivity analysis restricted to resting LVOTO, which was independent of provocation. Sixth, detailed hemodynamic and pharmacologic conditions at the time of follow-up TTE, including blood pressure, heart rate, hemoglobin, volume status, dehydration, and relevant medication exposure, were not consistently available. Because these factors can directly influence dynamic LVOT gradients, residual variability in the post-TAVR endpoint cannot be excluded. Seventh, pre-TAVR DLi-LVOTO was derived from a single resting PLAX view without Doppler integration and therefore may not fully capture the hemodynamic complexity of three-dimensional LV geometry or provoked obstruction; however, this simplicity enhances its practicality and scalability in routine practice. Finally, post-TAVR LVOTO was defined as a Doppler-based echocardiographic endpoint, and the present study did not establish whether this finding represented persistent, treatment-requiring, or clinically consequential obstruction. Its association with clinical outcomes such as hemodynamic compromise, rehospitalization, or mortality was not evaluated. Future studies with standardized follow-up and prospective outcome ascertainment are needed to clarify the clinical significance of this imaging phenotype.

In conclusion, in patients with severe AS undergoing TAVR, an HCM-trained DL model applied to pre-TAVR PLAX TTE was independently associated with Doppler-defined post-TAVR LVOTO, including in patients without pre-existing obstruction. These findings suggest that DLi-LVOTO may reflect a shared PLAX-based obstructive imaging phenotype associated with susceptibility to LVOT gradient elevation after TAVR. Further prospective multi-center studies with standardized follow-up and clinical outcome assessment are needed to establish its physiological and clinical significance.

## Author contributions

Jee-Woong Choi: Conceptualization, Data curation, Formal analysis, Investigation, Visualization, Writing – original draft, Writing – review & editing.

Jiesuck Park: Conceptualization, Data curation, Formal analysis, Investigation, Visualization, Writing – original draft, Writing – review & editing.

Yeonyee E. Yoon: Conceptualization, Funding acquisition, Project administration, Resources, Supervision, Writing – review & editing.

Jiyeon Kim: Methodology, Software, Validation, Writing – review & editing.

Jaeik Jeon: Methodology, Software, Validation, Writing – review & editing.

Yeonggul Jang: Methodology, Software, Writing – review & editing.

Seung-Ah Lee: Methodology, Software, Writing – review & editing.

Minjung Bak: Investigation, Writing – review & editing.

Hong-Mi Choi: Investigation, Writing – review & editing.

In-Chang Hwang: Investigation, Writing – review & editing.

Goo-Yeong Cho: Investigation, Writing – review & editing.

## Data availability

The data underlying this study cannot be made publicly available due to ethical restrictions set by the IRB of the study institution; i.e., public availability would compromise patient confidentiality and participant privacy. Please contact the corresponding author to request the minimal anonymized dataset. Researchers with additional inquiries about the deep learning model developed in this study are also encouraged to reach out to the corresponding author.

